# Association between Dysphagia and Symptoms of Depression and Anxiety after Ischemic Stroke

**DOI:** 10.1101/2023.08.23.23294519

**Authors:** Anel Karisik, Benjamin Dejakum, Kurt Moelgg, Silvia Komarek, Thomas Toell, Lukas Mayer-Suess, Raimund Pechlaner, Stefanie Kostner, Simon Sollereder, Sophia Kiechl, Sonja Rossi, Gudrun Schoenherr, Wilfried Lang, Stefan Kiechl, Michael Knoflach, Christian Boehme, the STROKE-CARD Registry study group.

## Abstract

**BACKGROUND:** Dysphagia after stroke is associated not only with poor outcome and higher mortality, but also with reduced quality of life and social isolation. We aimed to investigate the potential relationship between swallowing impairment and symptoms of anxiety and depression after ischemic stroke.

**METHODS:** Consecutive patients with ischemic stroke participating in the prospective STROKE-CARD Registry study at the study center Innsbruck, Austria from 2020 to 2022 were assessed for presence of dysphagia on hospital admission (clinical swallowing assessment) as well as for persistence until hospital discharge and a 3-month follow-up (SINGER Independency Index). Symptoms of anxiety and depression were recorded using Beck Depression Inventory (BDI) and Hospital Anxiety and Depression Scale (HADS) at 3-month follow-up.

**RESULTS:** Of 648 patients (36.6% female), 19.3% had dysphagia at hospital admission which persisted in 14.8% (hospital discharge) and 6.8% (3-month follow-up). With the presence or duration of dysphagia (no dysphagia, dysphagia at baseline, discharge or at 3 months), score points (mean±SD) increased in the BDI (7.9±6.7, 12.5±8.7, 13.5±9.0, 16.5±10.2), HADS-D (4.4±3.7, 7.1±4.2, 7.7±4.4, 9.8±4.3) and HADS-A (4.4±3.5, 5.4±3.6, 6.0±3.6, 7.0±3.6). In linear regression analysis adjusting for age, sex and functional disability, BDI and HADS-D but not HADS-A scores were significantly higher in patients with dysphagia when compared to those without dysphagia at baseline or who recovered to discharge and follow-up. Moreover, patients with swallowing impairment were more likely to receive antidepressants, antipsychotics or benzodiazepines at discharge and 3-month follow-up.

**CONCLUSIONS:** Dysphagia after stroke is common and severely affects psychosocial functioning of individuals. Our results highlight swallowing impairment as an independent predictor of depressive symptoms after stroke.

## INTRODUCTION

Advances in modern healthcare such as implementation of stroke unit care and reperfusion therapies have increased survival among individuals suffering from ischemic strokes.^1,2^ Despite this progress, a considerable proportion of patients face a wide range of complications and persisting impairments related to cerebral ischemia.^3^ Post-stroke dysphagia in particular, affecting 21 – 32% of patients after ischemic stroke, carries major complications, including increased risk of aspiration, pneumonia, and mortality.^4-9^ Moreover, from a psychosocial perspective, social life-impairing factors such as reduced quality of life and social isolation emerge in stroke patients with dysphagia, indicating the burden of swallowing impairment experienced by individuals in their daily functioning.^9^

Furthermore, oropharyngeal dysphagia, irrespective of etiology, appears to be related to affective symptoms such as anxiety and depression.^10^ While prior research examined the relation between dysphagia and affective symptoms in patients with head and neck cancer or Parkinson’s disease, recent findings indicate that individuals affected by stroke-related dysphagia might be more likely to develop post-stroke depression during hospital stay.^10,11^ Yet, post-stroke dysphagia differs from the chronic medical conditions above, as it has the potential to improve swallowing function after a period of time.

Our study aims to investigate the potential link between post-stroke dysphagia and symptoms of depression and anxiety in a large representative ischemic stroke cohort of the STROKE-CARD Registry within the first 3 months after onset.

## METHODS

### Study population

The post-stroke disease management program STROKE-CARD was implemented as a standard of care in December 2020 by the Department of Neurology at the Medical University of Innsbruck and the St. John of God Hospital in Vienna, Austria, after the STROKE-CARD study (NCT02156778) demonstrated efficacy of this program by reducing the cumulative risk of cardiovascular events and improving health-related quality of life.^12^ All patients with high-risk transient ischemic attack TIA (ABCD_2_ score ≥ 4) or ischemic stroke living in the catchment area of the respective centers receive a 3- and 12-month in-person follow-up visit to identify post-stroke complications, assess the patient’s need for care services, and support guideline-based secondary prevention, including lifestyle modifications and achievement of recommended target values. Additionally, besides general measures such as marital status, employment, need for support services and level of care (care levels according to the Austrian care support plan which provides financial aid for social care at home), clinical measures of stroke severity (National Institutes of Health Stroke Scale NIHSS^13^), disability (modified Rankin Scale mRS^14^), dependency (Scores of Independence for Neurologic and Geriatric Rehabilitation SINGER^15^) and quality of life (European Quality of Life 5 Dimensions 3 Level EQ-5D-3L^16^) are evaluated, along with comorbidities such as depression and anxiety disorder (measured by Beck Depression Inventory BDI^17^ and Hospital Anxiety and Depression Scale HADS^18^). For those signing an appropriate informed consent, data from baseline (i.e. hospitalization) as well as the 3- and 12-month follow-up visits is prospectively collected in the STROKE-CARD Registry.

For the present analysis, data from the study center Innsbruck was used. The Department of Neurology of the Medical University of Innsbruck with its stroke unit is the primary referral hospital for approximately 650.000 inhabitants and the secondary referral hospital for further 100.000 inhabitants of the county. Between December 2020 and October 2022 1076 patients fulfilled the inclusion criteria for the STROKE-CARD Registry. Of those 749 patients (69.6%) signed the informed consent during hospital stay or at the 3-month follow-up visit. After excluding 101 patients presenting with either (a) TIA (tissue-based definition) (n = 85), (b) pre-existing dysphagia (n = 3), and (c) early dropout of the study (n = 13), a total of 648 patients remained for the present analysis.

### Swallowing Assessment

The initial assessment of swallowing function was conducted through a review of clinical records. At the study center every stroke patient is screened for swallowing difficulties by experienced personnel as part of routine clinical care. Diagnosis of dysphagia was established via clinical examination of swallowing function by language and speech therapists, or using further instrumental diagnostics such as fiberoptic endoscopic evaluation of swallowing (FEES) if necessary. Based on the findings of these evaluations, along with dietary recommendations, the patient cohort was categorized in a dysphagia and a non-dysphagia group. The duration of swallowing difficulties was evaluated using logopedic protocols and the “eating/drinking” sub-item of the SINGER score^15^, which is utilized by therapists to determine whether swallowing issues were still present at hospital discharge or at the 3-month follow-up. Scores of 0-5 can be obtained, with a score of 5 indicating the absence of a swallowing problem. Patients with a SINGER score < 5 were classified as having persistent swallowing issues.

### Evaluation of Depression and Anxiety

History of depression was defined as intake of antidepressive medication at hospital admission or when documented in the digital health record. Presence of depressive and anxiety symptoms was assessed using the questionnaires (Beck Depression Inventory - BDI and Hospital Anxiety and Depression Scale - HADS) and the health-related quality of life measure EQ-5D-3L for the anxiety and depression domain after three months.^16-18^ All patients fill out the questionnaires either at home shortly before the 3-month follow-up or directly at the follow-up visit. Both psychiatric questionnaires (BDI and HADS) are validated screening instruments in the context of stroke.^19,20^ The Beck Depression Inventory includes 21 items with an overall maximum score of 63 points, while the Hospital Anxiety and Depression Scale provides 7 items each for anxiety and depression (14 items in total) with an overall maximum score of 42 points.^17,18^ Additionally, medications prescribed for depression and anxiety were recorded both at the time of hospital discharge and after three months. Psychiatric medication groups prescribed included antipsychotics (typical such as haloperidol as well as atypical such as risperidon, olanzapine, and others), antidepressants (tetracyclics, selective serotonin reuptake inhibitors, selective and non-selective serotonin norepinephrine reuptake inhibitors, noradrenergic and specific serotonergic antidepressant), and benzodiazepines (including imidazopyridin-derivates).

### Statistical Analysis

Age, sex and other characteristics were analyzed by presenting descriptive statistics such as mean and standard deviation or median and interquartile range for continuous and ordinal data, and percentage frequencies for binary data. To compare the groups with and without dysphagia, Pearson’s chi-square was used for binary and nominal variables, while the Mann-Whitney U test was used for ordinal or continuous, non-normally distributed data. Normal distribution of continuous data was tested using the Shapiro-Wilk test and the Kolmogorov-Smirnov test. To account for multiple testing in univariate comparisons, Bonferroni adjustment was applied. In cases of missing data, the adjusted sample was indicated for each variable. Additionally, multivariate linear regression was conducted to explore the independent association between dysphagia and depression/anxiety. The significance level was set at p < 0.05. Statistical analyses were performed using SPSS (Version 27.0.1.0; IBM Corporation, Armonk, New York).

## RESULTS

Baseline analyses were conducted on a cohort of 648 patients diagnosed with acute ischemic stroke, whose mean age was 71.0 years (±13.3 [SD]; range 23-101), with 36.6% of the study population being female.

### Post-Stroke Dysphagia

During the initial hospitalization period, dysphagia was observed in 19.3% (125 out of 648) of individuals. When limiting the analysis to the residents of Innsbruck and surroundings where our hospital serves as primary care center for stroke patients, a similar proportion showed dysphagia at baseline (18.9%, 82 out of 435). A comparison between patients with and without dysphagia revealed that those with swallowing difficulties were older (74.7 ± 12.0 vs. 70.2 ± 13.5 years, p < 0.001) and had a higher prevalence of atrial fibrillation (31.2% vs. 17.6%, p < 0.001) and diabetes (30.4% vs. 18.7%, p = 0.004). In addition, they experienced more severe strokes (NIHSS 7 [4-14] vs. 2 [1-4], p < 0.001) and greater disability (mRS 4 [3-5] vs. 2 [2-3], p < 0.001), as demonstrated in Table 1.

**Table 1.**
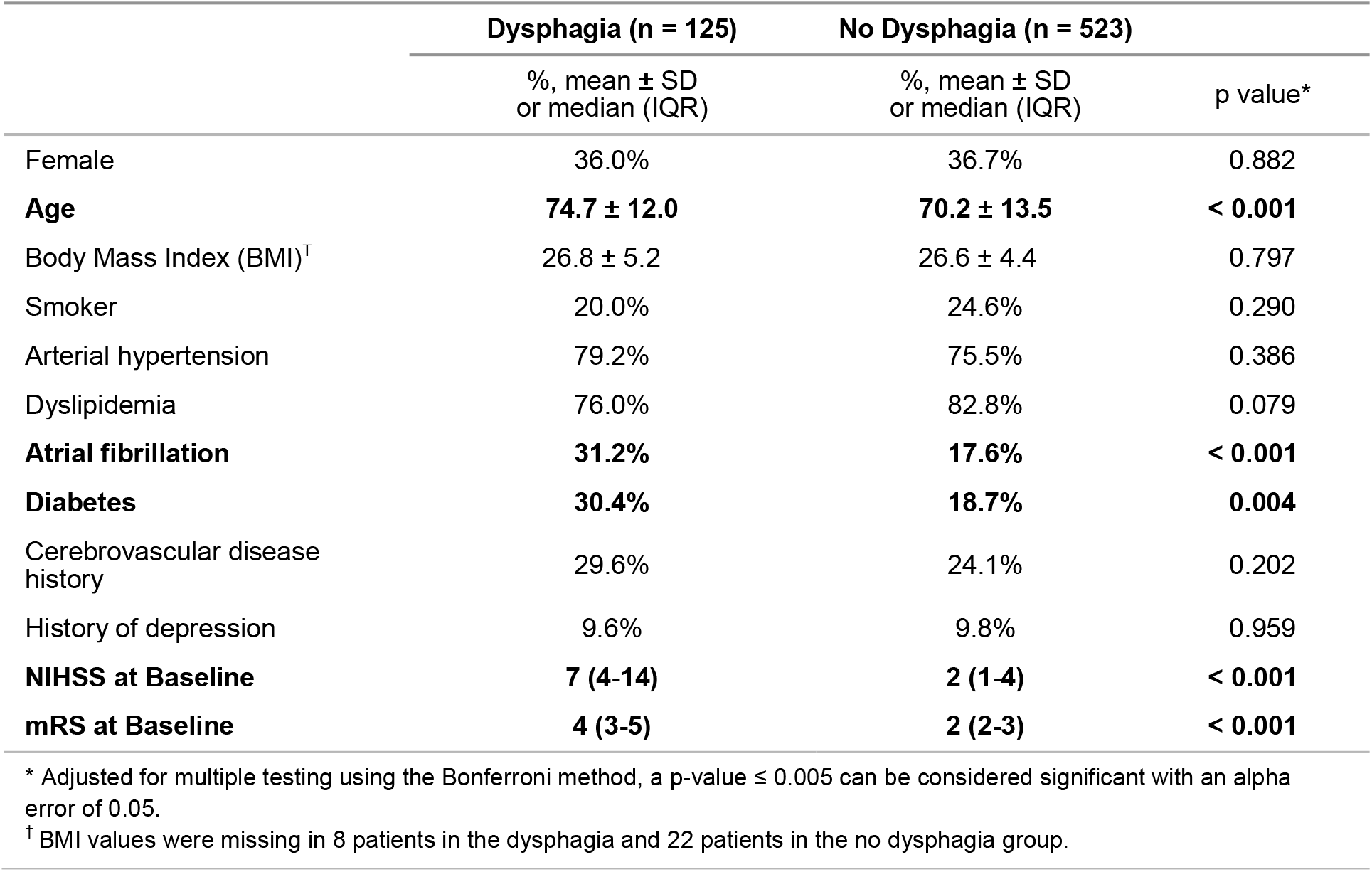
Baseline Characteristics of patients with or without Dysphagia.

|  | Dysphagia (n = 125) | No Dysphagia (n = 523) |  |
| --- | --- | --- | --- |
| | %, mean ± SD<br>or median (IQR) | %, mean ± SD<br>or median (IQR) | p value* |
| Female | 36.0% | 36.7% | 0.882 |
| **Age** | **74.7 ± 12.0** | **70.2 ± 13.5** | **< 0.001** |
| Body Mass Index (BMI)<sup>†</sup> | 26.8 ± 5.2 | 26.6 ± 4.4 | 0.797 |
| Smoker | 20.0% | 24.6% | 0.290 |
| Arterial hypertension | 79.2% | 75.5% | 0.386 |
| Dyslipidemia | 76.0% | 82.8% | 0.079 |
| **Atrial fibrillation** | **31.2%** | **17.6%** | **< 0.001** |
| **Diabetes** | **30.4%** | **18.7%** | **0.004** |
| Cerebrovascular disease<br>history | 29.6% | 24.1% | 0.202 |
| History of depression | 9.6% | 9.8% | 0.959 |
| **NIHSS at Baseline** | **7 (4-14)** | **2 (1-4)** | **< 0.001** |
| **mRS at Baseline** | **4 (3-5)** | **2 (2-3)** | **< 0.001** |

\* Adjusted for multiple testing using the Bonferroni method, a p-value $\leq 0.005$ can be considered significant with an alpha error of 0.05.
<sup>†</sup> BMI values were missing in 8 patients in the dysphagia and 22 patients in the no dysphagia group.

**Table 2.** Frequency of Dysphagia at baseline, discharge and 3-month follow-up.

|  | n | Dysphagia<br>% (n) |
| --- | --- | --- |
| **Baseline** | 648 | 19.3% (125) |
| **Discharge** | 648 | 14.8% (96) |
| **Follow-up** | 527 | 6.8% (36) |

At hospital discharge (median 8 [1-80] days after admission), 14.8% (96 out of 648) of patients had persistent dysphagia, as indicated by the SINGER score. Patients with dysphagia experienced longer hospital stays (15.6 ± 10.2 vs. 9.9 ± 6.0, p < 0.001) and were more likely to be discharged to a rehabilitation center (56.1% vs. 20.3%, p < 0.001) or a nursing home (7.3% vs. 1.9%, p = 0.001). Furthermore, they exhibited higher scores for NIHSS (3 [1-5] vs. 0 [0-1], p < 0.001) and mRS (3 [2-4] vs. 1 [1-2], p < 0.001) at the time of discharge.

At three month follow-up (median 3 [1-6] months after discharge), 121 of 648 patients (18.7%) were lost to follow-up, 37 of 125 (29.6%) in the dysphagia and 84 of 523 (16.1%) in the non-dysphagia group. Reasons were death in 18 patients (13 in the dysphagia and 5 in the non-dysphagia group), withdrawal of consent in 32 patients (9 in the dysphagia and 23 in the non-dysphagia group), visit via telephone-call in 20 patients (3 in the dysphagia and 17 in the non-dysphagia group) or missed appointments in 51 patients (12 in the dysphagia and 39 in the non-dysphagia group). 88 out of 125 patients who initially presented with dysphagia were examined in a follow-up assessment. Among those, 36 patients reported ongoing swallowing difficulties, which accounted for 6.8% of the follow-up cohort (36 out of 527) and 40.9% of initially dysphagic patients (36 out of 88). A similar proportion of stroke patients residing in Innsbruck and surroundings (5.1%, 22 out of 435) showed swallowing problems at follow-up.

Patients with initial dysphagia were more likely to live in nursing homes (20.2% vs. 2.8%, p < 0.001), require more external support services (32.9% vs. 9.9%, p < 0.001), had higher levels of care (46.8% vs. 15.6%, p = 0.001), and exhibit higher scores on the NIHSS (2 [1-4] vs. 0 [0-1], p < 0.001) and mRS (3 [2-3] vs. 1 [0-2], p < 0.001) scales after three months. Furthermore, health-related quality of life at 3 months was significantly lower in patients with initial dysphagia at baseline (59.1±23.6 vs. 71.9±21.3, p < 0.001) and follow-up (67.1±19.2 vs. 76.9±17.2, p < 0.001). Patients with dysphagia had a reduced BMI at 3 months (median drop in BMI 1.2, p < 0.001) whereas those without dysphagia remained the same (median drop 0.01, p = 0.630).

### Depression and Anxiety

At the 3-month follow-up, 389 questionnaires (73.8%) were collected, including 50 from patients with initial dysphagia (56.2%). Those patients which completed follow-up and submitted questionnaires were younger than those without questionnaires (68.5±13.1 vs. 75.0±12.7 years, p < 0.001) and had lower NIHSS (0 [0-1] vs. 1 [0-2], p < 0.001) and mRS (1 [0-2] vs. 2 [1-3], p < 0.001) scores at follow-up.

Patients with initial swallowing impairment had higher scores in all questionnaires after three months when compared with those without dysphagia, which was statistically significant for the HADS-D (7.1 ± 4.2 vs. 4.4 ± 3.7, p < 0.001) and the BDI (12.5 ± 8.7 vs. 7.9 ± 6.7, p < 0.001), but not for the HADS-A (5.4 ± 3.6 vs. 3.3 ± 3.5, p = 0.051) (Table 3).

**Table 3.** Mean Scores in HADS-A, HADS-D and BDI at the 3-month follow-up dependent on presence or persistence of dysphagia.

|  | No Dysphagia at Baseline<br>n = 339 | Dysphagia at Baseline<br>n = 50 | Dysphagia at Discharge<br>n = 39 | Dysphagia at Follow-up<br>n = 17 |
| --- | --- | --- | --- | --- |
| **HADS-A (mean ± SD)** | **4.4 ± 3.5** | **5.4 ± 3.6** | **6.0 ± 3.6** | **7.0 ± 3.6** |
| unadjusted p value | --- | 0.051 | 0.006 | 0.003 |
| adjusted p value | --- | 0.664 | 0.220 | 0.137 |
| **HADS-D (mean ± SD)** | **4.4 ± 3.7** | **7.1 ± 4.2** | **7.7 ± 4.4** | **9.8 ± 4.3** |
| unadjusted p value | --- | < 0.001 | < 0.001 | < 0.001 |
| adjusted p value | --- | 0.025 | 0.006 | < 0.001 |
| **BDI (mean ± SD)** | **7.9 ± 6.7** | **12.5 ± 8.7** | **13.5 ± 9.0** | **16.5 ± 10.2** |
| unadjusted p value | --- | < 0.001 | < 0.001 | < 0.001 |
| adjusted p value | --- | 0.040 | 0.018 | 0.005 |

P-values for the difference to patients without dysphagia at baseline or who recovered to discharge and follow-up (unadjusted and adjusted for age, sex, diabetes and mRS at 3 months using linear regression analysis)

There were no significant differences in mean scores of HADS-A, HADS-D, and BDI between women and men (data not shown). Subgroup analysis revealed that individuals had higher scores in all questionnaires based on the duration of swallowing dysfunction (from hospital discharge to the 3-month follow-up) (Table 3).

In a linear regression analysis that accounted for age, sex, diabetes and functional disability (mRS) at 3 months, patients who initially had dysphagia had significantly higher HADS-D (p = 0.025) and BDI (p = 0.040) scores at 3 months compared to those without dysphagia. However, there was no significant difference in HADS-A (p = 0.664) scores at 3 months. The same results were observed when comparing patients who had persistent dysphagia until hospital discharge (n = 39) or follow-up (n = 17) with those who never had dysphagia or recovered from dysphagia until hospital discharge (n = 349) or follow-up (n = 371), respectively (Table 3).

These findings fit to the results from EQ-5D-3L subdomain “anxiety/depression” that were available in 92.1% of the study population. While 45.6% of patients with initial dysphagia reported anxiety/depression after 3 months, only 24.6% of patients without dysphagia did. Figure 1 illustrates the distribution of the reported symptoms in patients with and without dysphagia.

**Fig. 1.**
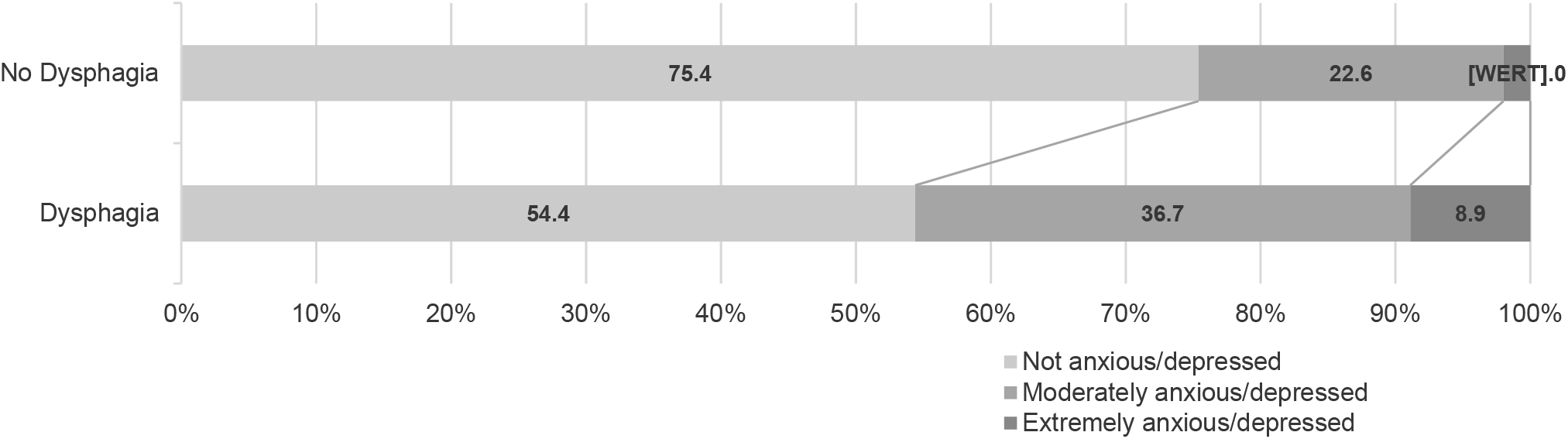
Comparison of EQ-5D-3L “anxiety/depression” sub-item between patients with or without initial dysphagia at the 3-month follow-up

The assessment of the newly initiated psychiatric medications (antipsychotics, antidepressants, benzodiazepines) prescribed at discharge and after 3 months indicated that patients with dysphagia had a higher frequency of prescriptions compared to those without dysphagia, both at discharge and after 3 months (Table 4). The prescription of antidepressants increased after 3 months, while prescriptions of antipsychotics and benzodiazepines decreased compared to discharge. Women had a significantly higher likelihood of receiving antidepressants at discharge and 3 months (13.5% vs. 6.3%; p = 0.004 and 22.8% vs. 9.6%; p < 0.001). At hospital admission, 9.7% of patients (63 out of 648) had pre-existing psychiatric medications (9.6% vs. 9.8% of patients with vs. without dysphagia, respectively), which all were excluded in the subsequent analysis.

**Table 4.**
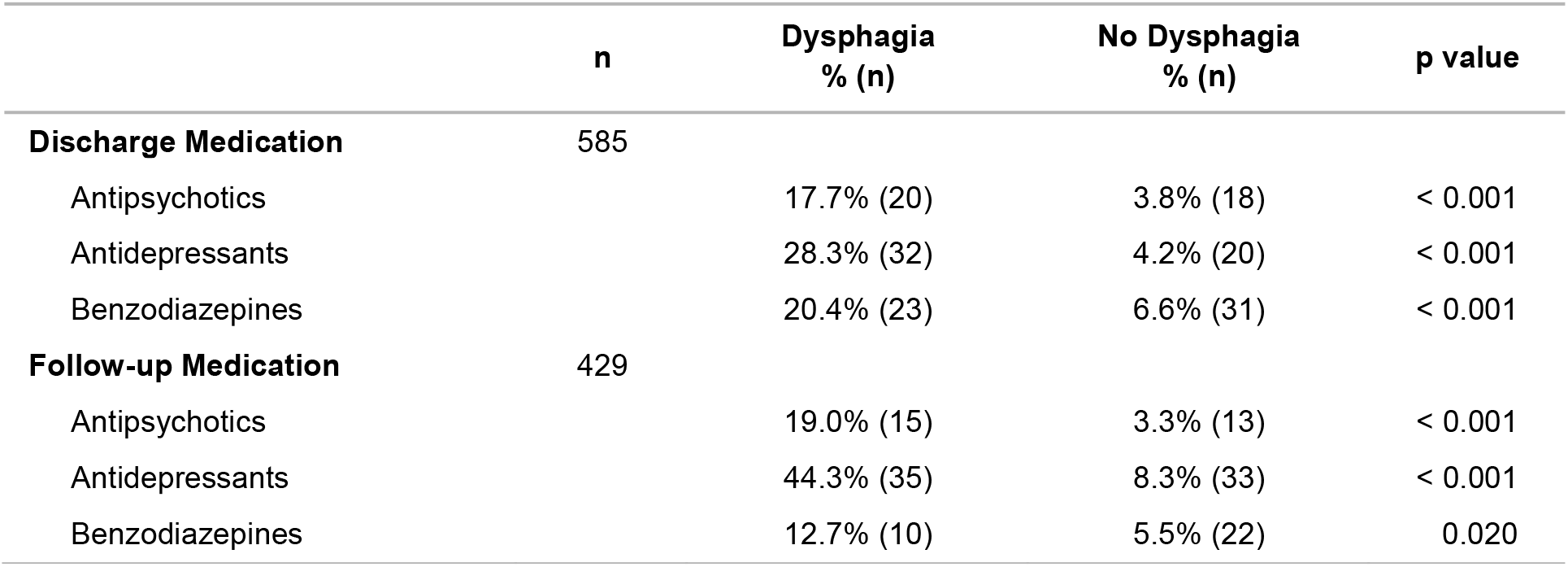
Prescription of novel psychiatric medications at hospital discharge and the 3-month follow-up.

## DISCUSSION

In the present analysis of the STROKE-CARD Registry study – a large representative cohort of consecutive ischemic stroke patients – we report the prevalence of post-stroke dysphagia in the acute phase and its recovery within the first 3 months as well as its association with symptoms of anxiety and depression.

To the best of our knowledge, this is the first study to report a clear association between post-stroke dysphagia and symptoms of depression and anxiety. The respective scores were higher with persistence of dysphagia and were associated with an increased prescription of Antipsychotics, Antidepressants or Benzodiazepines. When adjusting for age, sex and general functional recovery only the association with symptoms of depression but not anxiety remained statistically significant.

Support for our observation comes from a recent analysis of administrative data demonstrating an increased incidence of in-hospital depression in patients with stroke and dysphagia.^11^ On the other hand, in a cohort of patients with in-hospital rehabilitation, those with post-stroke depression were more likely to have dysphagia.^21^ Yet, both studies did not adjust for potential confounders.

The etiology and underlying principles of post-stroke dysphagia and post-stroke depression have been thoroughly studied in the past, including its pathogenesis and pathophysiology. Both conditions were linked to a variety of brain regions, yet no clear lesion pattern can be established.^22,23^ An alternative possible pathophysiologic connection between both entities is a disturbance of the neuronal network connectivity that has been linked to both – post-stroke dysphagia and post-stroke depression – separately.^10,23,24^

There are numerous reports that link oropharyngeal dysphagia as a consequence of other conditions, like Parkinson’s disease or orophyaryngeal cancer to affective symptoms (systematically reviewed^10^). Therefore, it can be assumed that not only neurobiological factors, but also psycho-social factors lead to an increase of depressive symptoms in patients with post-stroke dysphagia. In fact, the inability to have a regular meal leads to a substantial loss of pleasure, self-esteem and social withdrawal.^25^ As social isolation has been identified as a major contributor to the development of post-stroke depression^26^, a possible relation between these two factors appears to be plausible.

Even though our data show a clear relationship between the duration of dysphagia and the severity of depressive symptoms, it is unclear if an appropriate treatment and management of dysphagia decreases mood disturbances. Only a small study found improved BDI scores in 25 patients with intensified dysphagia exercise program, compared to 25 age and sex matched controls on standard care.^27^ Yet, as post-stroke depression is a relevant problem leading to substantial adverse effects on functional recovery, quality of life, and mortality^28,29^, intensified treatment and management of dysphagia might offer additional benefits in post-stroke depression treatment.

A second important finding in our large representative cohort of patients with ischemic stroke is the prevalence and prognosis of post-stroke dysphagia. In our sample - that also included very mild strokes - post-stroke dysphagia still was a prevalent condition affecting approximately 20% of patients in the acute phase that persisted in almost 80% at hospital discharge and 40% at 3-month follow-up. The frequency of dysphagia in the acute phase of stroke is similar with other studies reporting frequencies ranging from 21% to 32%^4-9^, but considerably lower than cohorts excluding patients with very mild strokes.^30^ There are diverging reports about recovery during follow-up with persistent dysphagia in the first few weeks of 51%^4^ or 17.5%^31^ and 2%^32^ or at least 50% (exact proportion of initially dysphagic patients unknown)^33^ at 3 or 6 months follow-up. When using videofluroscopic assessment swallowing difficulties were found in 80% of patients during follow-up.^33^ These differences in reported prognosis of dysphagia among studies can be explained by variations in assessment of dysphagia and patient populations (also including patients with hemorrhagic stroke^31-33^). Our study has utilized the “eating and drinking” item of the SINGER Independency Index, a well-established, standardized tool that effectively examines a patient’s independency status during neurorehabilitation, involving a multidisciplinary approach.^15^

The particular strength of our study is the inclusion of consecutive ischemic stroke patients, including the whole spectrum of ischemic stroke. Furthermore, we assessed symptoms of depression and anxiety using different scales validated in the context of stroke. Even though our assessment of dysphagia at discharge and follow-up (SINGER) does not replace standardized swallowing examinations, the evaluation was done according to the standardized assessment of a well-established rehabilitation scale by experienced personnel.^15^

When interpreting our results, one has to keep in mind that the BDI and HADS questionnaires were completed in 73.8% of the whole cohort. Those providing the questionnaires tended to be younger and less severely affected by their stroke. However, this selection bias is likely to attenuate our observed effect. In addition, a sensitivity analysis of the EQ-5D-3L “anxiety/depression” sub-item that was available in more than 90% of our studied cohort also strongly links dysphagia to affective symptoms. Lastly, the high level of prescribed psychiatric medications – that was available in every single patient – also indicates a strong correlation. A further limitation is the rate of patients lost to follow-up (30% in the dysphagia group vs. 16% in the non-dysphagia group). Additionally, concerning prescribed benzodiazepines, we can’t rule out the fact that some patients receive benzodiazepines solely as a sleeping aid instead of a depressive disorder.

## CONCLUSION

Post-stroke dysphagia is a prevalent complication among patients with ischemic stroke and has a major impact on psychosocial functioning, due to its association with depressive symptoms. Moreover, our results highlight dysphagia as an independent predictor in the development of depressive symptoms after ischemic stroke. Screening for post-stroke depression is crucial in all stroke patients, especially in those with dysphagia.

## Data Availability

We hereby confirm the availability of all data covered in the manuscript.

## Non-standard Abbreviations and Acronyms

BDI: Beck Depression Inventory
BMI: Body-Mass-Index
EQ-5D-3L: European Quality of Life 5 Dimensions 3 Level
FEES: Fiberoptic Endoscopic Evaluation of Swallowing
HADS: Hospital Anxiety and Depression Scale
mRS: modified Rankin Scale
NIHSS: National Institutes of Health Stroke Scale
SINGER: Scores of Independence for Neurologic and Geriatric Rehabilitation
TIA: Transient Ischemic Attack

## ACKNOWLEDGMENTS

We would like to acknowledge the efforts of the entire language and speech therapy team of the Department of Neurology, Innsbruck, for the great support.

## DISCLOSURES

The authors report no disclosures relevant to this research.

### The STROKE-CARD Registry Study Group

Gregor Broessner, Michael Eller, Julia Ferrari, Ton Hanel, Katharina Kaltseis, Theresa Köhler, Stefan Krebs, Florian Krismer, Christoph Mueller, Wolfgang Nachbauer, Anna Neuner, Anja Perfler, Theresa Schneider, Christine Span.

## REFERENCES

1. Campbell BCV, De Silva DA, Macleod MR, Coutts SB, Schwamm LH, Davis SM, Donnan GA. Ischaemic stroke. Nat Rev Dis Primers. 2019;5:70. doi: 10.1038/s41572-019-0118-8

2. Langhorne P, Ramachandra S, Collaboration SUT. Organised inpatient (stroke unit) care for stroke: network meta-analysis. Cochrane Database Syst Rev. 2020;4:CD000197. doi: 10.1002/14651858.CD000197.pub4

3. Winstein CJ, Stein J, Arena R, Bates B, Cherney LR, Cramer SC, Deruyter F, Eng JJ, Fisher B, Harvey RL, et al. Guidelines for Adult Stroke Rehabilitation and Recovery: A Guideline for Healthcare Professionals From the American Heart Association/American Stroke Association. Stroke. 2016;47:e98–e169. doi: 10.1161/STR.0000000000000098

4. Arnold M, Liesirova K, Broeg-Morvay A, Meisterernst J, Schlager M, Mono ML, El-Koussy M, Kägi G, Jung S, Sarikaya H. Dysphagia in Acute Stroke: Incidence, Burden and Impact on Clinical Outcome. PLoS One. 2016;11:e0148424. doi: 10.1371/journal.pone.0148424

5. Henke C, Foerch C, Lapa S. Early Screening Parameters for Dysphagia in Acute Ischemic Stroke. Cerebrovasc Dis. 2017;44:285–290. doi: 10.1159/000480123

6. Beharry A, Michel P, Faouzi M, Kuntzer T, Schweizer V, Diserens K. Predictive Factors of Swallowing Disorders and Bronchopneumonia in Acute Ischemic Stroke. J Stroke Cerebrovasc Dis. 2019;28:2148–2154. doi: 10.1016/j.jstrokecerebrovasdis.2019.04.025

7. De Cock E, Batens K, Hemelsoet D, Boon P, Oostra K, De Herdt V. Dysphagia, dysarthria and aphasia following a first acute ischaemic stroke: incidence and associated factors. Eur J Neurol. 2020;27:2014–2021. doi: 10.1111/ene.14385

8. Ko N, Lee HH, Sohn MK, Kim DY, Shin YI, Oh GJ, Lee YS, Joo MC, Lee SY, Song MK, et al. Status Of Dysphagia After Ischemic Stroke: A Korean Nationwide Study. Arch Phys Med Rehabil. 2021. doi: 10.1016/j.apmr.2021.07.788

9. Cohen DL, Roffe C, Beavan J, Blackett B, Fairfield CA, Hamdy S, Havard D, McFarlane M, McLauglin C, Randall M, et al. Post-stroke dysphagia: A review and design considerations for future trials. Int J Stroke. 2016;11:399–411. doi: 10.1177/1747493016639057

10. Verdonschot RJCG, Baijens LWJ, Vanbelle S, van de Kolk I, Kremer B, Leue C. Affective symptoms in patients with oropharyngeal dysphagia: A systematic review. J Psychosom Res. 2017;97:102–110. doi: 10.1016/j.jpsychores.2017.04.006

11. Horn J, Simpson KN, Simpson AN, Bonilha LF, Bonilha HS. Incidence of Poststroke Depression in Patients With Poststroke Dysphagia. Am J Speech Lang Pathol. 2022;31:1836–1844. doi: 10.1044/2022_AJSLP-21-00346

12. Willeit P, Toell T, Boehme C, Krebs S, Mayer L, Lang C, Seekircher L, Tschiderer L, Willeit K, Rumpold G, et al. STROKE-CARD care to prevent cardiovascular events and improve quality of life after acute ischaemic stroke or TIA: A randomised clinical trial. EClinicalMedicine. 2020;25:100476. doi: 10.1016/j.eclinm.2020.100476

13. Brott T, Adams HP, Olinger CP, Marler JR, Barsan WG, Biller J, Spilker J, Holleran R, Eberle R, Hertzberg V. Measurements of acute cerebral infarction: a clinical examination scale. Stroke. 1989;20:864–870. doi: 10.1161/01.str.20.7.864

14. Quinn TJ, Dawson J, Walters MR, Lees KR. Reliability of the modified Rankin Scale: a systematic review. Stroke. 2009;40:3393–3395. doi: 10.1161/STROKEAHA.109.557256

15. Gerdes N, Funke UN, Schüwer U, Themann P, Pfeiffer G, Meffert C. [“Scores of Independence for Neurologic and Geriatric Rehabilitation (SINGER)” - development and validation of a new assessment instrument]. Rehabilitation (Stuttg). 2012;51:289–299. doi: 10.1055/s-0031-1287805

16. Szende A, Janssen B, Cabases J. Self-Reported Population Health: An International Perspective based on EQ-5D. In; 2014.

17. Beck AT, Ward CH, Mendelson M, Mock J, Erbaugh J. An inventory for measuring depression. Arch Gen Psychiatry. 1961;4:561–571. doi: 10.1001/archpsyc.1961.01710120031004

18. Snaith RP. The Hospital Anxiety And Depression Scale. Health Qual Life Outcomes. 2003;1:29. doi: 10.1186/1477-7525-1-29

19. Aben I, Verhey F, Lousberg R, Lodder J, Honig A. Validity of the beck depression inventory, hospital anxiety and depression scale, SCL-90, and hamilton depression rating scale as screening instruments for depression in stroke patients. Psychosomatics. 2002;43:386–393. doi: 10.1176/appi.psy.43.5.386

20. Berg A, Lönnqvist J, Palomäki H, Kaste M. Assessment of depression after stroke: a comparison of different screening instruments. Stroke. 2009;40:523–529. doi: 10.1161/STROKEAHA.108.527705

21. Harris GM, Collins-McNeil J, Yang Q, Nguyen VQ, Hirsch MA, Rhoads CF, Guerrier T, Thomas JG, Pugh TM, Hamm D, et al. Depression and Functional Status Among African American Stroke Survivors in Inpatient Rehabilitation. J Stroke Cerebrovasc Dis. 2017;26:116–124. doi: 10.1016/j.jstrokecerebrovasdis.2016.08.039

22. Jones CA, Colletti CM, Ding MC. Post-stroke Dysphagia: Recent Insights and Unanswered Questions. Curr Neurol Neurosci Rep. 2020;20:61. doi: 10.1007/s11910-020-01081-z

23. Wang Z, Shi Y, Liu F, Jia N, Gao J, Pang X, Deng F. Diversiform Etiologies for Post-stroke Depression. Front Psychiatry. 2018;9:761. doi: 10.3389/fpsyt.2018.00761

24. Dum RP, Levinthal DJ, Strick PL. Motor, cognitive, and affective areas of the cerebral cortex influence the adrenal medulla. Proc Natl Acad Sci U S A. 2016;113:9922–9927. doi: 10.1073/pnas.1605044113

25. Ekberg O, Hamdy S, Woisard V, Wuttge-Hannig A, Ortega P. Social and psychological burden of dysphagia: its impact on diagnosis and treatment. Dysphagia. 2002;17:139–146. doi: 10.1007/s00455-001-0113-5

26. Sarkar A, Sarmah D, Datta A, Kaur H, Jagtap P, Raut S, Shah B, Singh U, Baidya F, Bohra M, et al. Post-stroke depression: Chaos to exposition. Brain Res Bull. 2021;168:74–88. doi: 10.1016/j.brainresbull.2020.12.012

27. Kang JH, Park RY, Lee SJ, Kim JY, Yoon SR, Jung KI. The effect of bedside exercise program on stroke patients with Dysphagia. Ann Rehabil Med. 2012;36:512–520. doi: 10.5535/arm.2012.36.4.512

28. Espárrago Llorca G, Castilla-Guerra L, Fernández Moreno Mc, Ruiz Doblado S, Jiménez Hernández MD. Post-stroke depression: an update. Neurologia. 2015;30:23–31. doi: 10.1016/j.nrl.2012.06.008

29. Towfighi A, Ovbiagele B, El Husseini N, Hackett ML, Jorge RE, Kissela BM, Mitchell PH, Skolarus LE, Whooley MA, Williams LS, et al. Poststroke Depression: A Scientific Statement for Healthcare Professionals From the American Heart Association/American Stroke Association. Stroke. 2017;48:e30–e43. doi: 10.1161/STR.0000000000000113

30. Losurdo A, Brunetti V, Broccolini A, Caliandro P, Frisullo G, Morosetti R, Pilato F, Profice P, Giannantoni NM, Sacchetti ML, et al. Dysphagia and Obstructive Sleep Apnea in Acute, First-Ever, Ischemic Stroke. J Stroke Cerebrovasc Dis. 2018;27:539–546. doi: 10.1016/j.jstrokecerebrovasdis.2017.09.051

31. Broadley S, Croser D, Cottrell J, Creevy M, Teo E, Yiu D, Pathi R, Taylor J, Thompson PD. Predictors of prolonged dysphagia following acute stroke. J Clin Neurosci. 2003;10:300–305. doi: 10.1016/s0967-5868(03)00022-5

32. Paciaroni M, Mazzotta G, Corea F, Caso V, Venti M, Milia P, Silvestrelli G, Palmerini F, Parnetti L, Gallai V. Dysphagia following Stroke. Eur Neurol. 2004;51:162–167. doi: 10.1159/000077663

33. Mann G, Hankey GJ, Cameron D. Swallowing function after stroke: prognosis and prognostic factors at 6 months. Stroke. 1999;30:744–748. doi: 10.1161/01.str.30.4.744

